# Genomic assessment of within-host population variation in *Neisseria gonorrhoeae*: Implications for gonorrhoea transmission

**DOI:** 10.1101/2020.08.24.20181230

**Authors:** Melinda M. Ashcroft, Eric P. F. Chow, Darren Y.J. Lee, Vesna De Petra, Maree Soumilas, Marlene Tschaepe, Lei Zhang, Danielle J. Ingle, George Taiaroa, Allison L. Hicks, Yonatan H. Grad, Benjamin P. Howden, Christopher K. Fairley, Deborah A. Williamson

## Abstract

**Objectives:** Mathematical modelling and genomic analyses are powerful methods for investigating the transmission dynamics of *Neisseria gonorrhoeae*, however, often make the implicit assumption that *N. gonorrhoeae* isolates at different anatomical sites within the same individual are the same strain.

**Methods:** In this study, two approaches were used to explore genetic diversity. First, we examined a collection of stored, clinical *N. gonorrhoeae* isolates sourced from multiple anatomical sites of single individuals attending a sexual health clinic in Melbourne from 2011-2019. Second, we obtained multiple colony picks from primary clinical samples from individuals attending a sexual health clinic in Melbourne from 2019-2020. Whole genome sequencing and a variety of bioinformatics approaches were used to determine both within-host and within-sample genetic diversity.

**Results:** Thirty-seven individuals were identified that had cultured *N. gonorrhoeae* from two or more anatomical sites (urogenital, anorectal, or oropharyngeal), with a final dataset of 105 isolates. In 35/37 (94.6%) individuals, infections were highly similar at the genetic level, with identical MLST and NG-MAST profiles. Pairwise comparisons of isolates within each individual indicated that the maximum within-host pairwise SNP distance was 13 SNPs (median = 1, IQR: 0-3). Notably, four distinct multi-individual phylogenetic clusters were identified, where the maximum pairwise SNP distance was 19 SNPs (median = 6, IQR = 2-11). Similarly, comparisons of isolates within each primary sample indicated that the maximum pairwise SNP distance was 8 SNPs (median = 2, IQR:1-3).

**Conclusions:** This study suggests that in most cases of multi-site infection, the same strain of *N. gonorrhoeae* causes the infection at each anatomical site. However, WGS data alone cannot differentiate between the same infecting strain or (re)infections from the same transmission network. These data guide recommendations regarding optimal bioinformatic approaches to infer genetic relatedness of *N. gonorrhoeae* and will help inform future studies of gonorrhoea transmission and epidemiology.

**KEY MESSAGES:**

- In most cases of multi-site infection, the same strain of *N. gonorrhoeae* causes infection at each anatomical site (urogenital, anorectal, and oropharyngeal) within an individual.
- It is not possible, using genomics alone, to determine if an infection is from a single individual or multiple individuals within the same sexual network.
- Current literature reports an array of different bioinformatic methods for describing genetic diversity, highlighting that SNP thresholds are not directly transferable between studies.

## INTRODUCTION

*Neisseria gonorrhoeae* is the causative agent of gonorrhoea, the second most common sexually transmitted infection (STI) worldwide. ^1^ In Australia, the incidence of gonorrhoea has increased rapidly in the mid-2010s from 66.9 per 100,000 population in 2014 to 125.5 per 100,000 population in 2018. ^2^ Further, multidrug-resistant (MDR) *N. gonorrhoeae* have emerged as a major clinical and public health challenge, both in Australia and globally. ^3 4^ Mathematical modelling and whole genome sequencing (WGS) are powerful methods for investigating the transmission dynamics of *N. gonorrhoeae* and can be used to guide research and public health interventions. ^5^ However, studies that use these approaches often make the implicit assumption that individuals with *N. gonorrhoeae* isolated from multiple anatomical sites (i.e. urogenital, oropharyngeal, anorectal) are infected with the same strain at all sites, or do not account for the fact that simultaneous infections with different strains can exist at the same site, ^6-8^ which may lead to incorrect conclusions about transmission.

WGS-based studies have utilised assessment of within-host genetic diversity to develop singlenucleotide polymorphism (SNP) thresholds for defining the likelihood of transmission of *N. gonorrhoeae* between individuals; for example, by generating a transmission nomogram ^9^ or by hierarchical single-linkage clustering. ^10^ To date, however, approaches to determine within-host SNP thresholds have differed across studies, ^9-16^ depending on the epidemiological and spatiotemporal nature of the datasets and the bioinformatic approaches used to estimate SNPs. Given the increasing use of genomic studies in informing public health responses to gonorrhoea, it is critical to understand the within-host strain variation of *N. gonorrhoeae*, and the robustness or limitations of methods that have been used in such studies. The primary aim of this study was to determine the within-host diversity of *N. gonorrhoeae*, both within and between anatomical sites. In addition, we suggest a bioinformatic framework for future epidemiological or population genetics studies of *N. gonorrhoeae*.

## METHODS

### Isolate selection, microbiological testing, and whole genome sequencing

To investigate within-host genetic diversity, a retrospective study was performed of individuals attending the Melbourne Sexual Health Centre (MSHC; the major publicly funded sexual health centre in Victoria, Australia) who had positive *N. gonorrhoeae* cultures available from multiple anatomical sites between January 1^st^, 2011 and November 30^th^, 2019. One hundred and eight isolates were obtained from 37 asymptomatic and symptomatic adult individuals (>16 years) from urogenital, oropharyngeal, and/or anorectal sites (Supplementary Dataset, Table S1). To assess diversity across anatomical sites within the same individual, single colonies of *N. gonorrhoeae* were selected from stored cultures from individuals who had *N. gonorrhoeae* isolated from at least two different anatomical sites on the same day (Figure 1a).

**Figure 1.**
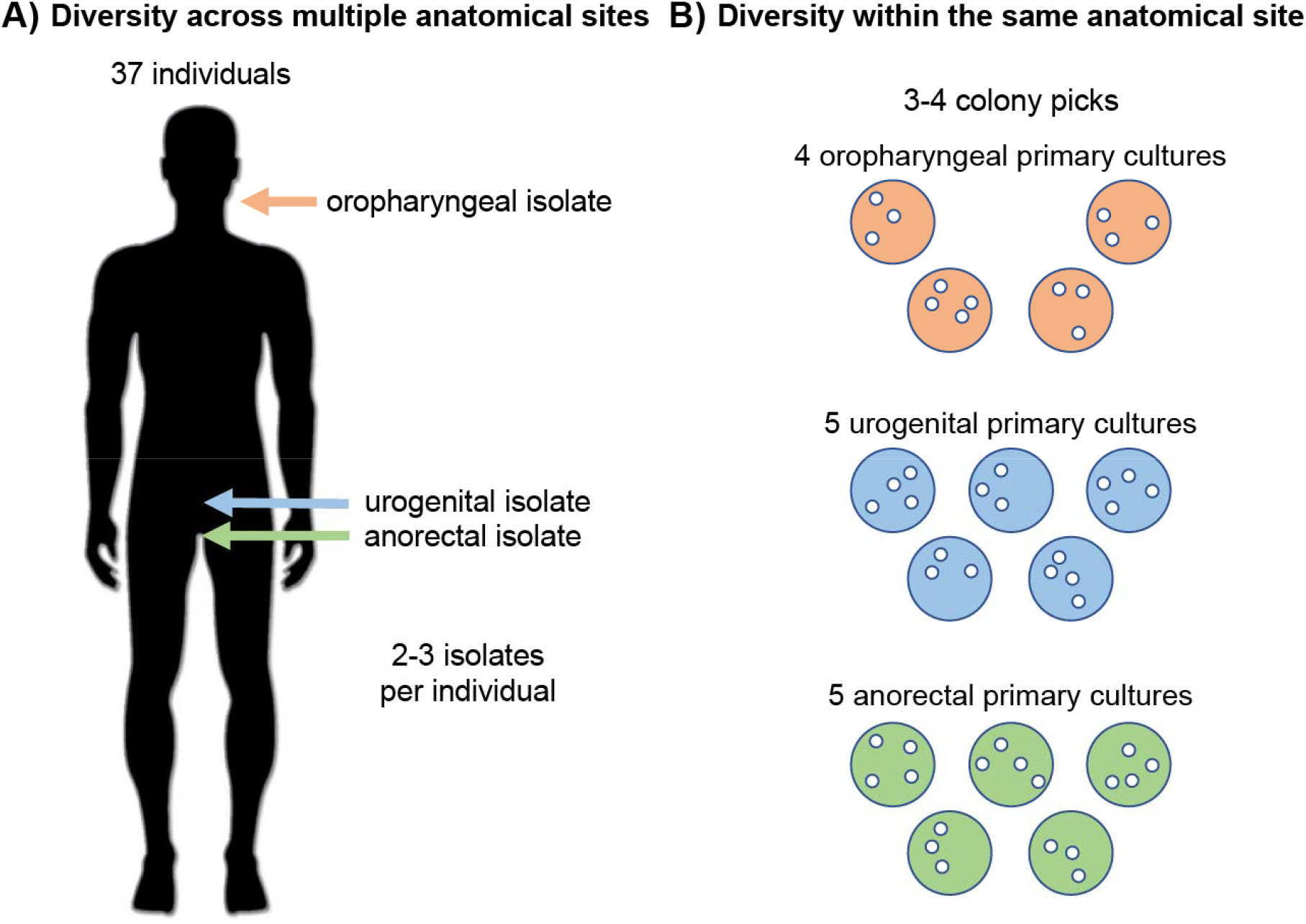
Approaches for assessing within-host and within-sample *N. gonorrhoeae* diversity. (A) Within-host diversity of isolates across at least two of three anatomical sites for 37 individuals. B) Within-site diversity of three to four colonies from four oropharyngeal, five urogenital and five anorectal primary cultures.

To investigate the genetic diversity present within the same anatomical site, 3-4 colony picks (selected on the basis of distinct gonococcal morphology) were taken from primary plates of a range of clinical samples (five anorectal swabs, four oropharyngeal swabs and five urogenital swabs from 13 individuals that were chosen at random) (Figure 1b and Supplementary Dataset, Table S2). Swabs were obtained from individuals attending the MSHC between November 25^th^, 2019 and March 4^th^, 2020. Isolates were confirmed as *N. gonorrhoeae* using a MALDI Biotyper (Bruker Daltonik, Bremen, Germany) and antimicrobial susceptibility testing (AST) was performed using agar breakpoint dilution as previously described. ^10^

### Whole genome sequencing, genome assembly and annotation

Genomic DNA (gDNA) was extracted from a single colony using the QIAsymphony™ DSP DNA Mini Kit (Qiagen) kit as the per manufacturer’s instructions. Isolates underwent WGS on an Illumina NextSeq 500 instrument with 150 bp or 300 bp paired-end reads using Illumina libraries and protocols. Raw Illumina sequence reads were processed using the *Nullarbor* pipeline (v2.0.20191007) ^17^ with default settings for each tool unless otherwise specified. Within this pipeline, reads were trimmed to remove adaptors and low-quality bases (Phred score <10) using Trimmomatic v0.39. ^18^ Kraken v1 using the MiniKraken 8Gb database (obtained 20/10/2017) was used to check for contamination. ^19^ Trimmed reads for each isolate were *de novo* assembled using Spades v3.14.0. ^20^ Genome annotation was performed using Prokka v1.14.2. ^21^ Multi-locus sequence typing (MLST) was performed using MLST v2.17.6, ^22^ updated 04/09/19 with the *Neisseria* PubMLST database ^23^ and multiantigen sequence types (NG-MAST) ^24^ were assigned using NGmaster v0.5.5. ^25^ Isolates were also typed *in silico* using the *N. gonorrhoeae* sequence typing for antimicrobial resistance (NG-STAR) ^26^ scheme implemented in pyngSTar. ^27^ Antimicrobial resistance genotype was correlated with phenotypic susceptibility data, with acquired resistance genes detected using Abricate v0.9.8 ^28^ and the NCBI Bacterial Antimicrobial Resistance Reference Gene Database (updated 18/03/18). ^29^

### Bioinformatic approaches used to define intra-patient genetic diversity

Current literature describing the within-host diversity of *N. gonorrhoeae* report an array of different bioinformatic methods, highlighting that SNP thresholds are not directly transferable across different datasets and studies (Supplementary Dataset, Table S3). Therefore, in this study, a variety of bioinformatic approaches were used to assess within-host *N. gonorrhoeae* pairwise SNP distances. Specifically, three metrics were examined: (i) choice of reference genome (applied to core-genome and individual-specific SNP alignments), (ii) masking of SNPs within repeats and mobile genetic elements (MGEs) and (iii) removing SNPs from recombinant regions (Supplementary Appendix, Fig S1). For metric (i), sequence reads were aligned to either the commonly used, antimicrobial-susceptible FA 1090 reference genome (Genbank accession: NC_002946; ML(ST)1899) or the complete genome AUSMDU00010541 (Genbank accession: NZ_CP045832; ST10899)), produced as part of a previous study, from Victoria, Australia, ^30^ to generate core-genome SNP alignments. For individual-specific SNP alignments, a single isolate from each individual was randomly chosen as a draft *de novo* reference genome and sequence reads from other isolates within that individual were aligned to the chosen reference. For all SNP calling, read alignment was done using bwa mem v0.7.17-r1188, ^31^ and SNPs were called using Freebayes v1.3.1, ^32^ requiring a minimum read coverage of 10, minimum base quality of 13% and 90% read concordance at a site as implemented within the Snippy pipeline. ^33^ The SNP density across each complete reference genome in 50 Kb segments was examined, which identified several clusters of high SNP density. Repeat regions for each reference genome were identified by aligning each reference genome to itself using the ‘--mask auto’ flag within snippy-core, which uses minimap2 ^34^ to create a self-homology map. Putative MGEs in each reference genome were identified using PHASTER ^35^ and IslandViewer 4. ^36^ For metric (ii), core-genome SNPs were then obtained from Snippy using the snippy-core flag, with SNPs in putative repeat and MGE regions masked using the --mask flag. The coordinates of each masked region from the AUSMDU00010541 reference genome can be found in Supplementary Dataset, Table S4. For metric (iii), the effect of recombination on the masked, core-genome SNP alignment for each reference genome was examined. Each SNP alignment was filtered for recombination using Gubbins v2.3.4 ^37^ with default settings in raxml mode, generating a phylogenetic tree with a generalised time-reversible model and gamma correction (GTRr). There was little difference in the size of the final, recombinant filtered, masked, core SNP alignments between the two reference genomes (9,050 and 9,491 SNPs against FA 1090 and AUSMDU00010541 respectively). Thus, we decided to use the contemporary, Victorian, Australia complete genome reference AUSMDU00010541 ^30^ for defining pairwise SNP distances and generating a phylogenetic tree.

Core-genome pairwise SNP distances generated with each complete genome (FA 1090 or AUSMDU00010541) were imported as a matrix into R v3.6.1. ^38^ These datasets were then converted into a long format, with intra-patient pairwise SNP distances analysed using harrietr. ^39^ Histograms and boxplots of pairwise SNP distances were plotted using ggplot2. ^40^ Hierarchical single-linkage cluster analysis was performed with the hclust function and clusters were filtered using a threshold of 13 SNPS (based on the maximum within-host diversity identified in this study) using the cutree function (R s*tats* package). The Mann–Whitney Rank sum test was used to compare non-normal distributions of pairwise SNP distances within and between hosts. P values were reported if below the α significance region (α = 0.05, two-sided test).

### Phylogenetic analyses

The core-genome SNP alignment (9,491 SNPs, representing a core-genome of 1,780,502 bp (81.87% of the AUSMDU00010541 reference genome)), where SNPs within repeats, MGEs and recombinant regions were masked, was used for phylogenetic analyses. This SNP alignment was used as input for IQ-Tree v1.6.12 ^41^ with the GTR+G4 model, constant sites (465,996, 513,232, 530,477, 475,038), ultrafast bootstrapping with 1000 replicates, and the SH-aLRT parameter with 1000 bootstrap replicates to infer a Maximum Likelihood (ML) phylogenetic tree. The resulting ML tree was loaded into R and visualised alongside genomic and phenotypic metadata using ggtree. ^42^

### Ethics

The study was approved by the Alfred Hospital Ethics Committee (399/19).

## RESULTS

### Effect of differing bioinformatic approaches on within-host and within-site diversity

Between 2011 and 2019, there were 37 individuals who had available *N. gonorrhoeae* isolates cultured from at least two anatomical sites, resulting in 108 isolates that underwent WGS. Sequence and assembly quality metrics were examined, with three isolates discarded due to contamination by other species, leaving a final collection of 105 isolates from 37 individuals (Supplementary Dataset, Table S1). Two individuals had urogenital and anorectal isolates, three individuals had urogenital and oropharyngeal isolates, one individual had anorectal and oropharyngeal isolates and 31 individuals had urogenital, anorectal, and oropharyngeal isolates. From these 105 isolates, two coregenome SNP alignments were generated. Alignments were generated against either *N. gonorrhoeae* genome FA1090, or a complete *N. gonorrhoeae* genome from Victoria, Australia (AUSMDU00010541). ^30^ This resulted in 19,522 and 18,934 core-genome SNPs across all 105 genomes, respectively. Several high-density SNP regions were identified in each core-genome SNP alignment (Supplementary Appendix, Fig S2). The impact of SNPs within repeat regions and MGEs was assessed, with masking of these regions resulting in 16,407 and 17,437 core-genome SNPs respectively. Additionally, SNPs within recombinant regions were assessed. Recombination events introduced 7,357 (44.84%) and 7,946 (45.57%) SNPs into each core-genome SNP alignment (Supplementary Appendix, Fig S3). Thus, filtering of recombination from the masked, core-genome SNP alignments resulted in 9,050 and 9,491 SNPs respectively.

Using the masked, recombination-filtered core SNP alignment against AUSMDU00010541 as a reference, there was minimal variation across anatomical sites within individuals. For 103/105 isolates (35/37 individuals; 94.6%), the maximum pairwise SNP distance across sites was 13 coregenome SNPs (median = 1, IQR: 0-3), consistent with within-host variation arising from one infection or as a result of reinfection by the same strain. Two individuals had one isolate each that had different MLST, NG-MAST and NG-STAR profiles and differed by a maximum of 87 and 778 core-genome SNPs respectively (individual 29 had a divergent oropharyngeal isolate and individual 17 had a divergent urogenital isolate (Supplementary Appendix, Note 1)), consistent with simultaneous infections of different strains at different anatomical sites (Figure 2a and 2b). Similarly, individual-specific core-SNP alignments indicated that for 35/37 individuals (94.6%), the maximum pairwise distance between isolates was 13 SNPs (median = 4, IQR: 3-5) (Figure 2c). However, two individuals had individual-specific core-SNP alignments of 956 and 5,128 SNPs respectively, reflective of the multiple simultaneous infections shown above, although with approximately 10-fold more SNPs detected using the individual-specific reference approach. As expected, the within-host diversity was significantly lower than the genetic diversity between individuals (range=0-2,649 SNPs, median=1,701, IQR: 888-1938, *P* < 0.0001).

**Figure 2.**
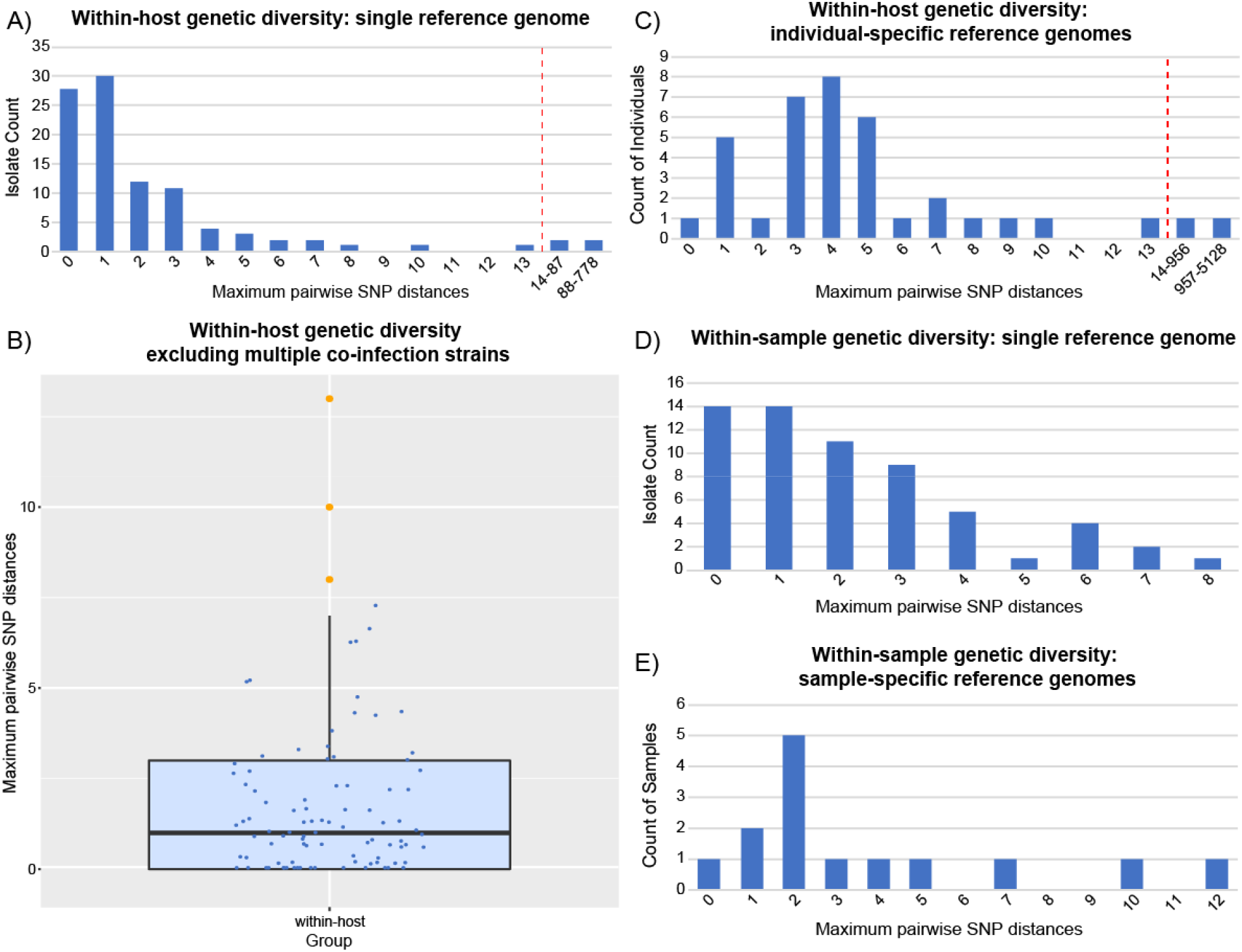
Within-host and within-sample genetic diversity. A) Within-host genetic diversity present across multiple anatomical sites using a single reference genome. B) Within-host genetic diversity excluding the multiple co-infection strains from individuals 17 and 29. Box plot indicates the median and inter-quartile range. Orange dots indicate outliers. C) Within-host genetic diversity using individual-specific reference genomes. D) Within-sample genetic diversity across multiple primary samples using a single reference genome. E) Within-sample genetic diversity using samplespecific reference genomes. Red dotted lines indicate maximum pairwise SNP distance for isolates to be designated as the same strain.

The within-sample diversity of *N. gonorrhoeae* isolates was also examined, with subculture and sequencing from 49 individual *N. gonorrhoeae* colonies obtained from primary samples (Supplementary Dataset, Table S2). Similar to the approach taken to assess within-host diversity, for within-sample isolates, a core-genome SNP alignment was generated against (i) AUSMDU00010541 (recombination filtered, with SNPs in repeats and MGEs masked), and (ii) a draft *de novo* reference from one isolate within that sample. The maximum pairwise SNP distance across all samples was 8 core-genome SNPs (median = 2, IQR: 1-3) (Figure 2d). Similarly, sample-specific core-SNP alignments indicated that pairwise SNP distances between isolates differed by a maximum of 12 SNPs (median = 2, IQR: 2-5) (Figure 2e, Supplementary Appendix, Fig S4 and Supplementary Appendix, Note 2). These results demonstrate that within-sample diversity for multiple colony picks is comparable to the within-host diversity identified across multiple anatomical sites. Notably however, the maximum pairwise SNP distances exceed those reported in a similar study, which showed limited genetic diversity (maximum pairwise SNP distance of 2 SNPs, median = 0) within multiple colony picks from single clinical samples. ^9^

### Sexual networks can confound SNP-based assessment of within-host diversity

Using a hierarchical single-linkage clustering method with a SNP threshold of 13 SNPs difference (based on the maximum within-host diversity defined in this study), four distinct multi-individual phylogenetic clusters were identified, the largest of which comprised isolates from eight individuals, obtained up to five years apart (Figure 3 and Supplementary Appendix, Fig S5). Comparisons of isolates within each cluster indicated that the maximum pairwise SNP distance within each cluster were similar to the within-host variation, ranging from 7 SNPs for Cluster 4 (median = 3, IQR: 2-4) to 19 SNPs (median = 6, IQR: 2-11) for Cluster 1 (Supplementary Dataset, Table S5).

**Figure 3.**
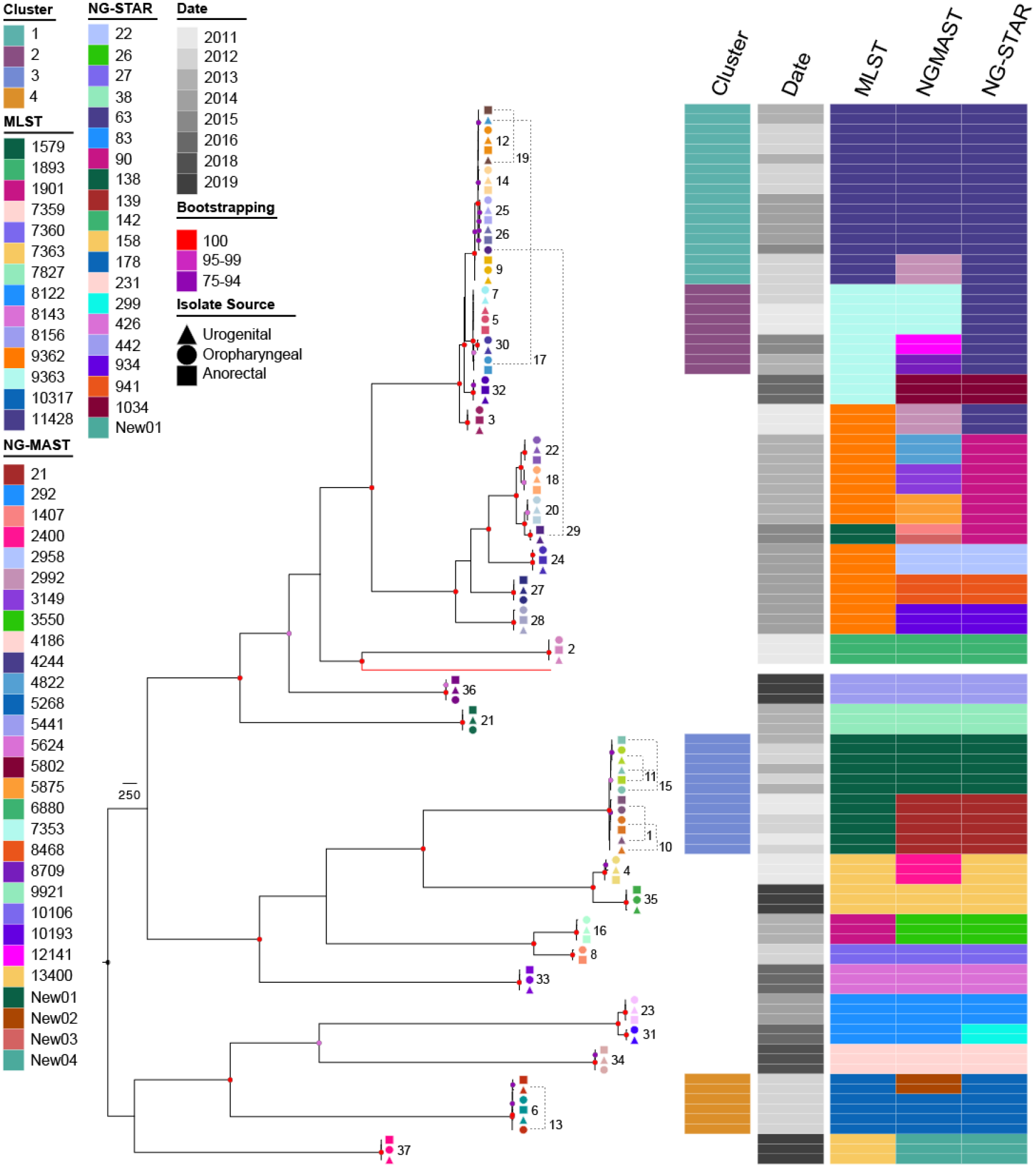
Phylogenetic relationship of *N. gonorrhoeae* isolates and individuals in this study. The mid-point rooted Maximum Likelihood phylogenetic tree was built from 9,491 core-genome SNPs against the AUSMDU00010541 reference genome (highlighted by red branch). Isolates from the same individual have been coloured the same to illustrate within-host diversity. Shapes represent infection site: urogenital (triangle), oropharyngeal (circle) and anorectal (square) isolates. Genomic metadata is shown adjacent to the tree. The date represents the year of isolation. For individuals with lines linking isolates, the number is displayed at the lower end of the joining line.

## DISCUSSION

Defining the within-host *N. gonorrhoeae* diversity is essential for understanding the transmission dynamics of *N. gonorrhoeae* between individuals, with SNP thresholds being a common method for defining isolates of the same strain or transmission event. In this study, using two different measures of genetic diversity, we found that isolates of *N. gonorrhoeae* from (i) the same sample and (ii) from multiple anatomical sites within an individual, were similar to isolates from different individuals if the strains of *N. gonorrhoeae* were from the same phylogenetic cluster. Excluding the two individuals with obvious co-infections, within-host isolates differed by a maximum of 13 SNPs at the core-genome level using a single reference genome, or by 13 SNPs using a within-host reference genome, similar to previous reports. ^9 10 15 16^ These data are similar to the maximum within-sample pairwise SNP distances of 8 SNPs at the core-genome level using a single reference genome, or 12 SNP using a within-sample reference genome. Similarly, there were four distinct clusters of isolates from multiple individuals, with identical MLSTs and maximum pairwise SNP distances in the same range as the maximum within-host variation, suggestive of a transmission network. Analysis of WGS data alone, however, cannot establish whether isolates at different anatomical sites represent infections from: (i) a single sexual encounter; (ii) transmission from the same person but from different sexual encounters, or (iii) from different people in the same sexual network. This is not possible because isolates within each of these samples are genetically indistinguishable. Nonetheless, this uncertainty can be partly mitigated by: (i) incorporating detailed epidemiological metadata into genomic assessments of transmission, ^10^ (ii) characterisation of the genetic diversity in the community in which the isolates are circulating, such as that shown in Williamson, *et al*. 2019, ^10^ (iii) longitudinal sampling of individuals where serial isolates are obtained over a defined time period from initial presentation, ^43^ or (iv) deep sequencing (samples sequenced to 500-1000x coverage) to identify mixed variants that would not be detected with standard (30-100x) sequence coverage (as has been shown in *Mycobacterium tuberculosis* ^44^).

Studies describing the within-host diversity of *N. gonorrhoeae* have employed different bioinformatic methods, ^9-16^ highlighting that SNP thresholds are not directly transferable across different datasets and studies. SNP-based thresholds are dataset-specific and are affected by numerous bioinformatic factors, demonstrated by our analysis in this study. These include: (i) how closely related the reference genome is; (ii) methods used to define the core-genome; (iii) whether SNPs within repeat regions and MGEs are filtered from the alignment; and (iv) whether SNPs within recombinant regions are filtered from the alignment. Additionally, the size of the coregenome (in general, smaller studies have larger core-alignments to the reference genome than larger studies ^45^) and the presence of outbreak-related strains may also affect SNP thresholds. ^46^

Based on the findings within this study, and in the context of other published literature, we propose the following as a bioinformatics framework for undertaking and reporting *N. gonorrhoeae* genomic and transmission studies: (i) inclusion of a closely related reference genome (which maximises the number of core genome sites and thus provides greater resolution to discriminate between closely related isolates ^45^); (ii) publication of detailed, reproducible and open-source methods for generating core-genome SNPs, including all tools, versions and parameters; (iii) filtering of SNPs from repeats and MGE regions, and (iv) where appropriate, filtering of SNPs from recombinant regions based on the size and composition of the dataset. Such standardisation will improve the comparability of results across studies, enable reproducibility and improve the accuracy of genomics-inferred “transmission events”. ^46^ However, with the diversity of tools, methods, and reference genomes available, some differences in SNP threshold are to be expected. Notably, inferences around transmission based on WGS data are increasingly being used in, for example forensic cases, contact tracing, and public health responses, ^47-49^ and it is thus imperative, that there is transparency and standardisation in the methods used for defining genetic diversity and SNP thresholds.

In conclusion, we suggest that in general, most *N. gonorrhoeae* isolates from an individual, within and between anatomical sites, are likely to be the same strain, supporting previous modelling studies that have made this assumption. Nonetheless, while we identified a maximum pairwise SNP distance to define isolates as the same strain, our data demonstrates that there is no SNP threshold that clearly delineates isolates as the same infecting strain as opposed to a reinfection by the same sexual network. This study highlights the need to integrate WGS and detailed epidemiological data to make accurate inferences regarding *N. gonorrhoeae* transmission dynamics, and ultimately enable public health interventions.

## Data Availability

Sequence data (reads) have been submitted to the National Center for Biotechnology Information (https://www.ncbi.nlm.nih.gov) under the
BioProject Accession: PRJNA520805. All supporting data have been provided within the article or supplementary data files.

## Acknowledgements

We thank Kerrie Stevens, Reference, Unusual Pathogens and Resistance (RUPR) section head and RUPR team at the Microbiological Diagnostic Unit, Public Health Laboratory (MDU PHL) for technical assistance. We also thank the staff of the molecular diagnostics and bioinformatics sections at the MDU PHL for antimicrobial susceptibility testing and sequencing of isolates and bioinformatics support.

## Funding

D.A.W, E.P.F.C and C.K.F are supported by an Investigator Grant from the National Health and Medical Research Council (NHMRC) of Australia (GNT1174555, GNT1172873 and GNT1172900, respectively).

## Notes

### Competing Interest Statement

The authors have declared no competing interest.

### Author Declarations

The study was approved by the Alfred Hospital Ethics Committee (399/19).

